# Early Life Safety Profiling of Gene Therapy for Spinal Muscular Atrophy: A Case Series Analysis

**DOI:** 10.1101/2024.11.04.24316576

**Authors:** Rebecca G. Spellman, Leillani L. Ha, Salomé Da Silva Duarte Lepez, Elizabeth A. Arruda, Emma Rodrigues, Kathryn J. Swoboda, Christiano R. R. Alves

## Abstract

The present study examines the safety profile of intravenous onasemnogene abeparvovec gene therapy in a real world setting, both alone or in combination with intrathecal antisense oligonucleotide nusinersen therapy in two cohorts of patients with spinal muscular atrophy (SMA). The first cohort included 8 presymptomatic infants treated solely with onasemnogene abeparvovec, while the second cohort comprised 6 symptomatic infants receiving onasemnogene abeparvovec and nusinersen co-therapy. All patients received the corticosteroid prednisolone coincident with gene therapy. Circulating alanine aminotransferase (ALT) and aspartate transaminase (AST) levels were measured to determine potential hepatoxicity, the primary focus of this study. Elevated ALT and AST levels, but no change in GGT levels were observed in 1 pre-symptomatic and 3 symptomatic patients post-treatment. However, all values normalized within three months of onasemnogene abeparvovec injection. Nusinersen treatment received previously or coincident with gene therapy did not impact the elevation of liver transaminases, which was transient. This study highlights the importance of early intervention with molecular treatments for SMA and indicates that prior or coincident treatment with nusinersen is unlikely to impact safety of onasemnogene apoparvovec and could theoretically improve clinical outcomes in symptomatic infants or in those with gene therapy delayed beyond the immediate neonatal period.

## Introduction

Molecular therapies have demonstrated remarkable clinical benefits when administered during the pre-symptomatic or early symptomatic stages of spinal muscular atrophy (SMA), significantly improving survival rates even among the most severely affected infants^1^. In 2016, nusinersen (Spinraza, Biogen) became the first molecular therapy approved for SMA, and numerous studies have since confirmed its efficacy and safety across various SMA populations^2,3^. In 2019, the U.S. Food and Drug Administration (FDA) approved onasemnogene abeparvovec (Zolgensma, Novartis/AVXS-101, AveXis), a gene therapy using an adeno-associated virus vector to deliver the SMN1 gene through a single intravenous injection in newborns and infants with SMA. Recent clinical trials have validated the effectiveness of onasemnogene abeparvovec in both pre-symptomatic and symptomatic infants, showing significant motor function improvements^4–6^. Over the past three years, this has largely become the treatment of choice for monotherapy in newborns with homozygous SMN1 deletion, regardless of SMN2 copy number. While gene therapy is a transformative treatment for SMA, varying adverse effects are observed: most commonly categorized as mild or moderate in severity these adverse events need to be closely monitored^4–6^. Despite limited data on the safety or long term outcomes of sequential overlapping or combinatorial treatment, there is an increasing number of infants receiving dual or sequential therapy with gene therapy and either nusinersen or risdiplam, an issue not adequately addressed yet in published clinical trials or studies. One of the potentially largest adverse impact on long term efficacy of gene therapy treatment in newborns with 2 SMN2 copies are the delays inherent in obtaining preauthorization, approval, and required preliminary safety testing prior to administration of AAV-9 mediated gene therapy. For a proportion of newborns, their catabolic state, variable transition to appropriate weight gain and social factors in the critical post-partum period, immediate dosing post birth may be neither ideal nor feasible. Thus adequate processes for rapid initiation of an alternative molecular therapy need to be standardized, and would ideally include overlapping coverage through the duration of the viral transduction process post dosing with AAV9-mediated intravenously delivered gene therapy.

The most common adverse effect reported with onasemnogene abeparvovec gene therapy is elevated liver enzyme levels, which can indicate acute liver injury^7,8^. In clinical practice, these elevated levels are usually managed effectively with co-administration of prednisolone, a corticosteroid hormone^8,9^. Clinical trials have demonstrated that initiating prednisolone 48 hours prior to gene therapy dosing and adjusting as needed for 2-3 months while closely monitoring liver function can successfully avoid serious medical consequences^10^. However, further longitudinal research is needed to fully understand the implications and risk of serious consequences at the extremes of age and weight not included in clinical trials.

Combinatorial therapies for SMA patients could be used more strategically to facilitate maximum rescue of at-risk motor neurons and other tissues^11^. The current approach in the clinical setting in the U.S. has been to choose one therapy, and then only consider adding another therapy if the first therapy proves to have insufficient benefit. In newborns identified by newborn screening, timing of gene therapy initiation is variable, but not infrequently delayed several weeks. In an infant with 2 SMN2 copies, this could lead to substantial progression of motor neuron denervation. Nusinersen and onasemnogene abeparvovec act via different molecular mechanisms and display different delivery effectiveness. Nusinersen loading has been associated with dramatic and persistent drop in neurofilament levels in 2 copy infants, and has a long half life that could persist through the gene therapy dosing and transduction process. Thus, in a presymptomatic clinical cohort of infants with 2 SMN2 copies, we hypothesize that sequential administration of nusinersen and onasemnogene abeparvovec could present a synergic effect in these vulnerable SMA infants, without the need to continue long term combinatorial therapy. In already symptomatic 2 copy infants, near simultaneous dosing at the earliest opportunity, as demonstrated in the cohort included here, could maximize survival of the greatest number of at risk motor neurons and help ensure maximal transduction efficacy. Here, we report for the first time a comprehensive safety dataset including the levels of circulating liver enzymes from SMA cases receiving treatment with only onasemnogene abeparvovec, and SMA cases receiving combinatorial therapy with nusinersen and onasemnogene abeparvovec in their first months of life.

## Methods

This is a longitudinal cohort study including a total of 14 SMA newborns or infants treated in the first months of life at the Massachusetts General Hospital (MGH), including 8 infants who received treatment with onasemnogene abeparvovec and 6 infants who received combinatorial treatment with nusinersen and then onasemnogene abeparvovec. All patients received prednisolone 48 hours prior to onasemnogene abeparvovec dosing and continued for 2-3 months post dosing as per standard of care. None of these patients were receiving risdiplam (Evrysdi, Roche). Of note, all patients receiving only onasemnogene abeparvovec were treated early in life at the pre-symptomatic stage, while all patients receiving co-therapy with nusinersen and onasemnogene abeparvovec in this study demonstrated clinical symptoms at dosing. The primary outcomes of this study are circulating alanine aminotransferase (ALT) and aspartate transaminase (AST) levels measured as biomarkers of potential hepatoxicity. In addition to the absolute values, the fold change from the baseline (i.e. measurements taken before injection) ALT and AST levels were also calculated for each individual patient and presented. Circulating gamma-glutamyl transferase, troponin T-hs Gen 5 (Troponin T), white blood counts, platelets and albumin levels were also determined as secondary outcomes along with any clinical adverse effect manifestation. Reference normal ranges were determined based on MGH laboratory references. Reference values from the Mayo Clinic and the University of California San Francisco (UCSF) Children’s are also provided. *SMN1* and *SMN2* copy numbers were determined using quantitative polymerase chain reaction as previously described^12^. All other outcomes were obtained from the clinical laboratory in association with research visits in the SPOT SMA Longitudinal Pediatric Data Repository (LPDR) housed within the Research Electronic Data Capture Web Application at the Newborn Screening Translational Research Network^13^. Written informed parental consent was obtained from all participants under Institutional Ethics Review Board at the MGH (protocol #2016-P000469). Distribution of age range and sex are presented in **Table 1**.

**Table 1.** Information of patients with SMA receiving onasemnogene abeparvovec early in life.

| Subject | Sex | Age range at dosing (days) |  |
| --- | --- | --- | --- |
| Onasemnogene abeparvovec monotherapy in presymptomatic newborns and infants up to ~ 6 months of age (n = 8) |  |  |  |
| 1 | F | 31-35 |  |
| 2 | M | 21-25 |  |
| 3 | F | 16-20 |  |
| 4 | F | 81-85 |  |
| 5 | F | 36-40 |  |
| 6 | M | 186-190 |  |
| 7 | F | 101-105 |  |
| 8 | M | 171-175 |  |
| Combinatorial therapy with onasemnogene abeparvovec (AVXS) and nusinersen in a symptomatic cohort (n = 6) |  |  |  |
| 9 | F | Nusinersen: 71-75 | AVXS: 936-940 |
| 10 | F | Nusinersen: 91-95 | AVXS: 96-100 |
| 11 | F | Nusinersen: 46-50 | AVXS: 141-145 |
| 12 | M | Nusinersen: 76-80 | AVXS: 81-85 |
| 13 | M | Nusinersen: 16-20 | AVXS: 26-30 |
| 14 | M | Nusinersen: 6-10 | AVXS: 96-100 |

## Results

We determined circulating ALT and AST levels in two cohorts of SMA patients receiving gene therapy early in life (**Table 1**). The first cohort included 8 SMA infants ages 20 – 190 days old treated at the pre-symptomatic stage. They received only onasemnogene abeparvovec treatment (**Figures 1A-H)**. The second cohort included 6 SMA infants and 1 child who received both nusinersen and onasemnogene abeparvovec (**Figures 2A-F**). Only 1 of 8 presymptomatic infants dosed with onasemnogene abeparvovec had elevated circulating ALT and AST levels, occurring within a few days post injection (**Figure 1B**), showing a 20-fold change in ALT and 9-fold change in AST (**Figure 3A-B, 3E-F**). This specific case was a subject with 3 *SMN2* copies. Importantly, increased ALT and AST levels returned to normal within 3 months after the injection (**Figure 1B, 3A-B, 3E-F**). For the second cohort including symptomatic patients with 2 *SMN2* copies, we observed 3 out of 6 cases with elevated circulating ALT and AST levels post onasemnogene abeparvovec injection (**Figure 2A-F**). Two patients showed similar changes, both displaying about a 4.5-fold increase in ALT and a 4-to-7-fold increase in AST. Another patient showed a 28-fold change in ALT and 6-fold change in AST from baseline (**Figure 3C-D, 3E-F**). Of note, this patient was also the oldest dosed patient. Increased ALT and AST levels also returned to normal levels around 3 months after the injection and were maintained at normal ranges during follow-up (**Figure 2A, 2C, 2E, 3C-D, 3E-F**). Taken together, these data closely parallel previously published clinical trial data demonstrating that some infants with SMA experience transient hepatotoxicity after gene therapy even when prednisolone is co-administered, but these effects are transient and typically return to normal by three months. Importantly, all enrolled patients had normal albumin levels and no indications of malnutrition or chronic liver disease (**Table 2** and **Supplementary Table 1**).

**Table 2.** Clinical characteristics of patients with SMA receiving onasemnogene abeparvovec early in life.

| Subject | SMN2<br>(copies) | GGT<br>(U/L) | Troponin T<br>(ng/L) | WBC<br>(K/uL) | Platelets<br>(K/uL) | Albumin<br>(g/dL) | HGB<br>(g/dL) | CK<br>(U/L) | CK-MB<br>(ng/mL) | No. of<br>Adverse<br>Events |
| --- | --- | --- | --- | --- | --- | --- | --- | --- | --- | --- |
| Onasemnogene abeparvovec monotherapy in presymptomatic newborns and infants up to ~ 6 months of age (n = 8) |  |  |  |  |  |  |  |  |  |  |
| 1 | 2 | 14-66 | *Not tested* | 6.58-13.16 | 346-497 | 3.7-4.8 | 10.5-13.2 | 125-618 | 8.5-32.8 | 9 (1 SAE) |
| 2 | 2 | 10 | <50 | 10.8-11.2 | 371-378 | 3.9-4.4 | 14.1-14.7 | *Not tested* | *Not tested* | 1 |
| 3 | 3 | 12-76 | 5 | 10.40-13.50 | 365-730 | 3.8-4.7 | 9.7-13.7 | 40-154 | 3.8-7.1 | 4 |
| 4 | 3 | 11-29 | 7-29 | 7.06-11.6 | 319-446 | 3.7-4.4 | 9.8-12.1 | 49-168 | 3.2-5.9 | 0 |
| 5 | 3 | 7-13 | <6-25 | 13.44-19.85 | 347-609 | 3.7-4.5 | 11.2-12 | 79-82 | *Not tested* | 10 |
| 6 | 4+ | 11-26 | 7-28 | 5.61-10.82 | 422-914 | 4.0-4.8 | 8.9-14.3 | 188 | 5.8 | 0 |
| 7 | 3 | 7-16 | 6-7 | 6.02-9.56 | 363-407 | 4.2-4.4 | 10.4-12 | 95-211 | 4.4-10.8 | 9 |
| 8 | 4+ | 18-34 | 8-21 | 3.96-7.48 | 181-333 | 4.3-4.6 | 11.5-12.7 | 229 | 10.8 | 1 |
| Combinatorial therapy with onasemnogene abeparvovec (AVXS) and nusinersen in a symptomatic cohort (n = 6) |  |  |  |  |  |  |  |  |  |  |
| 9 | 2 | 7-72 | 19-61 | 7.23-13.11 | 156-541 | 3.5-4.7 | 9.5-14.8 | 54-74 | 5.2-7.9 | 8 (2 SAEs) |
| 10 | 2 | 5-50 | 33-82 | 9.74-14.72 | 318-541 | 4.1-4.9 | 10.6-12.9 | 106-248 | 9.9 | 5 (1 SAE) |
| 11 | 2 | 6-18 | 60 | 6.75-13.50 | 316-476 | 4.2-4.7 | 10.9-13.2 | 105-150 | 15.4 | 16 |
| 12 | 2 | 8-46 | 41-90 | 5.71-10.85 | 289-423 | 4.0-5.1 | 11-14 | 126-148 | 15.5-24.6 | 8 |
| 13 | 2 | 4-22 | 42-61 | 7.07-11.9 | 267-799 | 3.3-4.7 | 9.4-13.8 | 160-141 | *Not tested* | 9 (3 SAEs) |
| 14 | 2 | 11-23 | 25-68 | 7.04-8.18 | 199-399 | 4.0-4.8 | 10.9-12 | *Not tested* | *Not tested* | 1 |

GGT: Gamma-glutamyl transferase; Troponin T: Troponin T-hs Gen 5; WBC; White blood cell count. SAE: severe adverse event

**Figure 1.**
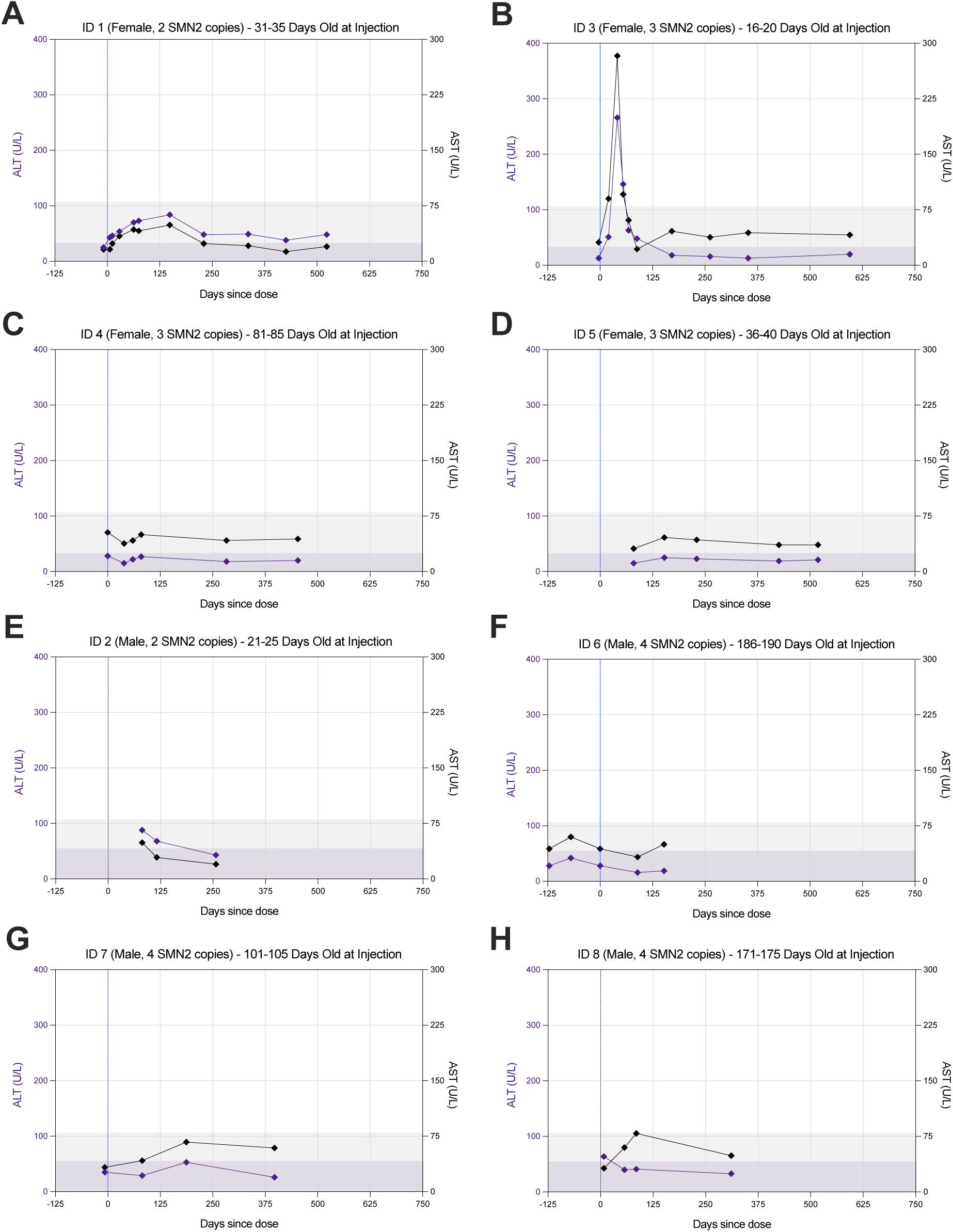
Circulating alanine aminotransferase (ALT) and aspartate transaminase (AST) levels in a cohort of SMA patients treated at the pre-symptomatic stage with onasemnogene abeparvovec. Blue line indicates the day of injection. Colored background area indicates normal concentrations expected in sex-matched controls.

**Figure 2.**
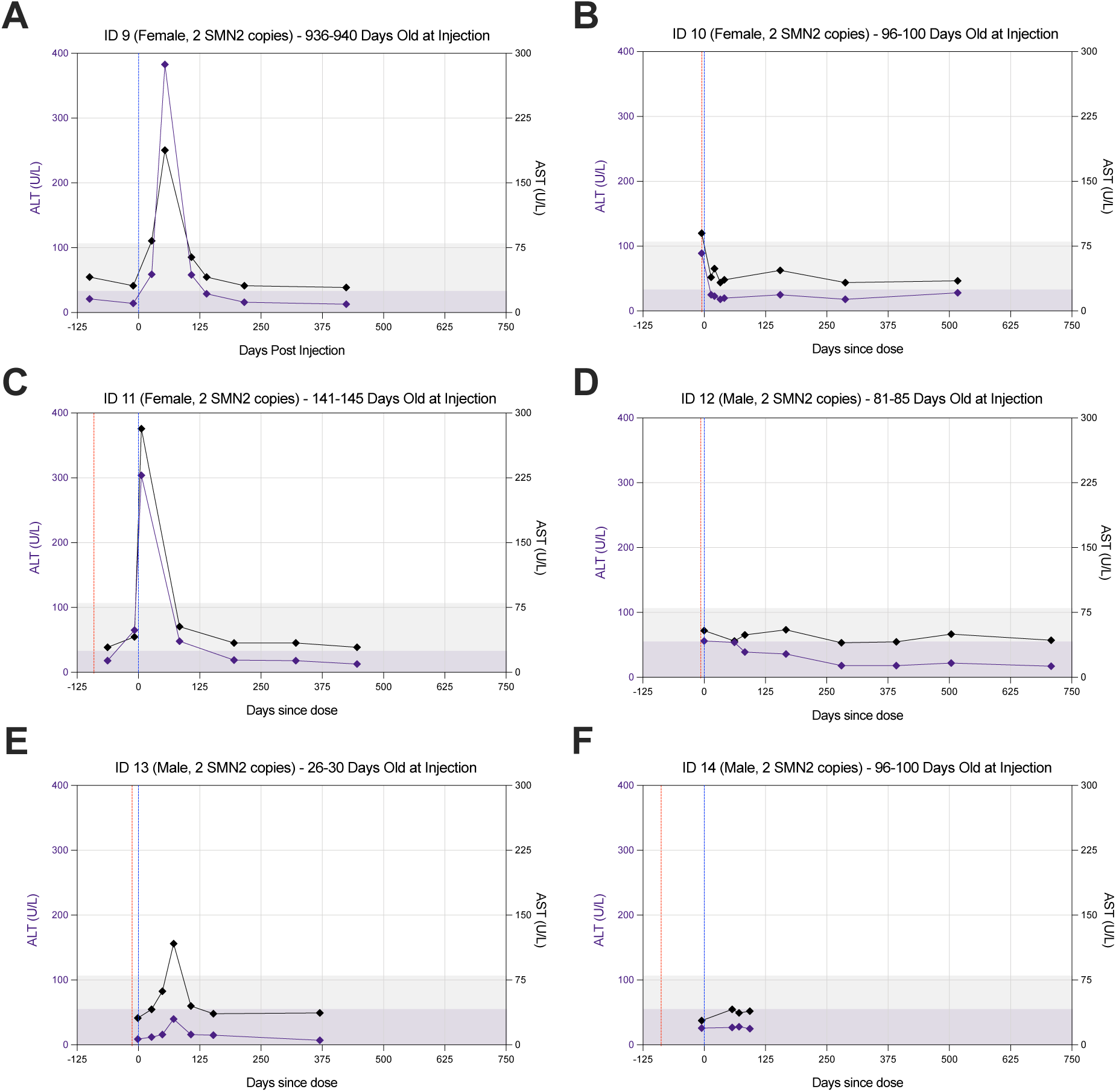
Circulating alanine aminotransferase (ALT) and aspartate transaminase (AST) levels in a cohort of SMA patients treated with nusinersen and onasemnogene abeparvovec. Blue line indicates the day of onasemnogene abeparvovec injection. Red line indicates the initial nusinersen dose for each case. Colored background area indicates normal ALT and AST concentrations expected in sex-matched controls. Note: Case #9 has initiated nusinersen treatment more than 125 days before the onasemnogene abeparvovec dose.

**Figure 3.**
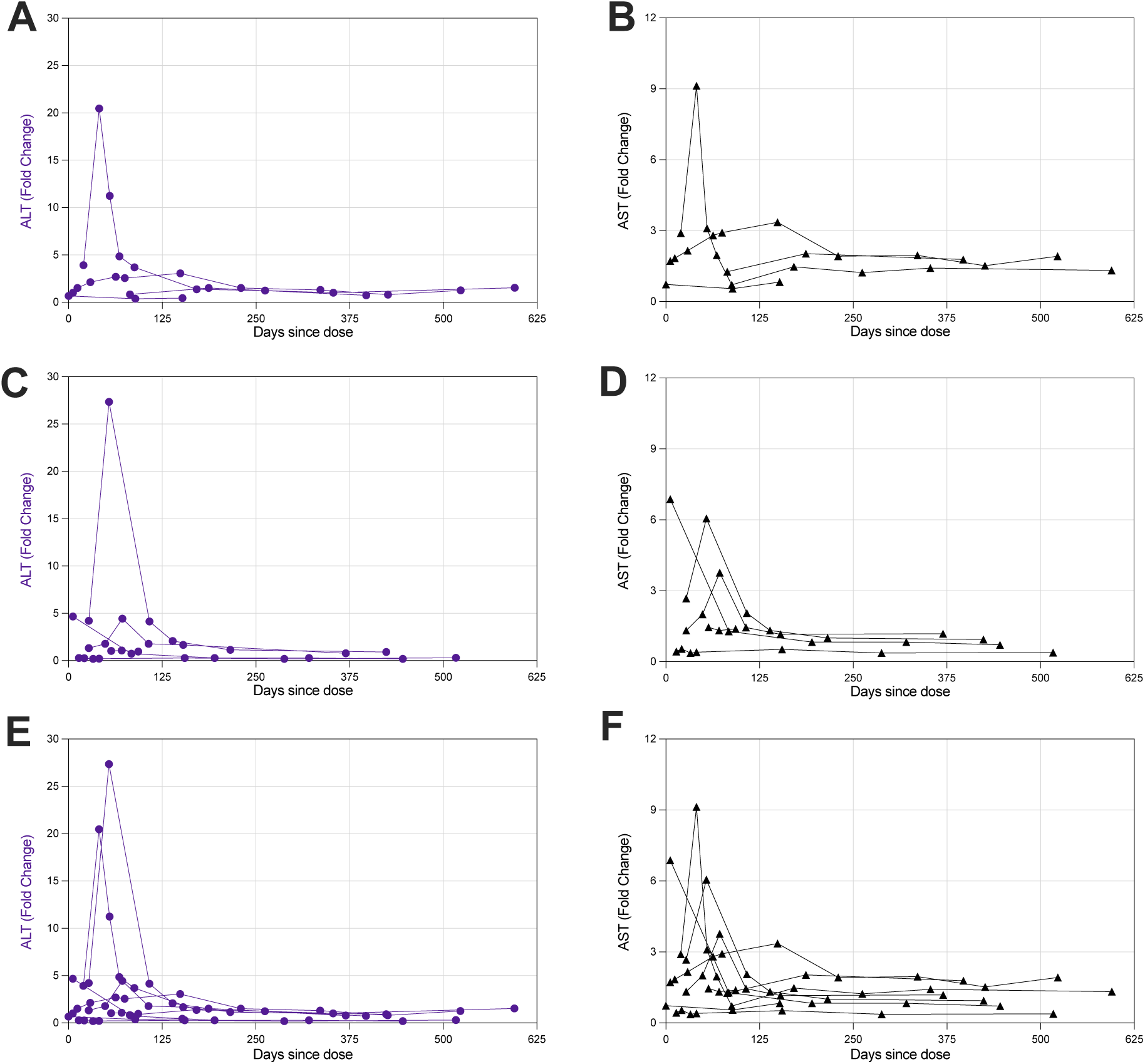
Fold change of circulating alanine aminotransferase (ALT) and aspartate transaminase (AST) levels in SMA patients treated with onasemnogene abeparvovec or with nusinersen and onasemnogene abeparvovec. **a-b**, Cohort receiving onasemnogene abeparvovec monotherapy. **c-d**, Cohort receiving nusinersen and onasemnogene abeparvovec co-therapy. **e-f**, Both cohorts displayed together in the same plot. AST and ALT were normalized based on the baseline (pre treatment) for each patient.

As complementary analysis, we observed increased levels of troponin T and platelets in most of the SMA infants and children (**Table 2** and **Supplementary File 1**), indicating potential myocardial injury and thrombocytosis. These increased levels are most likely part of the natural course of SMA pathophysiology since they were not observed as a direct consequence of any therapy. Other measurements such as circulating white blood counts and platelets also demonstrated some specific values out of normal range during this longitudinal follow-up study, but did not clearly correlate with treatment (**Table 2** and **Supplementary File 1**). Moreover, we observed multiple mild or moderate clinical events during the follow up of this study, but only 4 patients treated with either gene therapy or co-therapy showed at least one severe adverse event related to a cardiac, respiratory, or nutritional issue (**Table 3**). These adverse events were managed in hospital visits. They were not clearly associated with treatment response, and were considered most likely related to the primary underlying diagnosis. Altogether, these findings validate previous evidence showing that patients with SMA need to be closely monitored after transformative gene therapy is administered in the real world clinical setting.

**Table 3.** Summary of adverse events (AEs) in patients with SMA receiving onasemnogene abeparvovec early in life.

|  | **Mild/Moderate** | **Severe/Hospital Visit Needed** | **Total** | **%** |
| --- | --- | --- | --- | --- |
| **Fever** | 8 | 1 | 9 | 11.4% |
| **GI** | 9 | --- | 9 | 11.4% |
| **Cardiac/Acute life threatening** | 1 | 1 | 2 | 2.5% |
| **Respiratory** | 6 | 3 | 9 | 11.4% |
| **Orthopedic** | 5 | --- | 5 | 6.3% |
| **Nutritional** | 1 | 1 | 2 | 2.5% |
| **Other** | 39 | 4 | 43 | 54.4% |
| **TOTAL AEs** | 69 | 7 | 79 |  |
| **%** | 87.3% | 8.9% |  |  |

## Discussion

Taking advantage of our longitudinal database (MGH SPOT SMA LPDR) that includes SMA cases treated early in life with onasemnogene abeparvovec, we were able to evaluate several markers related to the safety of gene therapy. Our findings indicate that some SMA patients can present hepatotoxicity within a few days after receiving the gene therapy. More drastic changes in ALT and AST values were notably observed in patients dosed with gene therapy at an older age in the second cohort in contrast to the first cohort whose age at dosing was younger on average, reemphasizing the importance of initiating therapy in 2 *SMN2* copy infants at the earliest opportunity. These effects are transient and return to normal levels within three months post injection. Corroborating these single center observations, a study combining data from 5 previous clinical trials has observed liver-related adverse events in 34% of SMA patients treated with gene therapy^7^. Variable liver response to the gene therapy was also clearly observed in the original trial establishing the single-dose gene therapy for SMA and emphasized the importance of co-treatment with oral prednisolone to mitigate immune-mediated hepatotoxicity^8^. Prednisolone doses can be adjusted when necessary in each specific SMA case^8^. Here, we observed a total of 4 SMA cases suggesting at least transient liver injury in spite of prednisolone treatment, but without any evident long-lasting consequences.

We report longitudinal safety data from 6 symptomatic SMA cases with only 2 *SMN2* copies that have been treated with co-therapy with onasemnogene abeparvovec and nusinersen. These patients are achieving motor milestones that were not expected in the natural history of the disease and the present data also suggest that side effects of these therapies are transient and can be controlled by closely monitoring each patient. These results demonstrate the safety for sequential and coincident nusinersen therapy during the induction process for gene therapy. Nusinersen treatment administered pre-gene therapy treatment theoretically provides an advantage over risdiplam, given its prolonged treatment effect following an initial loading period. However, we did not examine the impact or safety of sequential or coincident treatment with risdiplam in this study, and thus our conclusions are limited. In line with our findings, another recent study including 2 SMA cases that received co-therapy with nusinersen and onasemnogene abeparvovec also reported no apparent increased burden of adverse effects^11^. This possibility of combining therapies for SMA patients has presented itself as particularly interesting for the SMA community. Based on our current data, we advocate for additional studies analyzing different outcomes in SMA patients receiving co-therapies and an open discussion about the advantages and disadvantages of combining these treatments. We hypothesize that there is limited to no benefit in continuing a second therapy once gene therapy has been delivered in a truly presymptomatic infant with 2 copies. However, there may be in an infant with some evident denervation and an emerging clinical phenotype. In our experience, progressive denervation can sometimes occur quite precipitously in a 2 copy infant between ordering and dosing gene therapy. Thus, biomarkers such as CMAP, MUNE, or robust serum biomarkers must be integrated into the current clinical paradigm since clinical exam and even functional motor tests remain insufficient for this purpose.

We observed increased troponin T and variable platelet levels in SMA patients, which are markers of increased risk for myocardial injury and thrombocytosis. While damage in motor neurons accounts for the earliest manifestations in SMA infants, evidence indicates that other tissues contribute to the SMA pathophysiology due to the direct effects of SMN deficiency in tissues other than motoneurons, or indirect effects due to the severe denervation status^14,15^. In this context, a multidisciplinary approach is critical to monitor and provide the best care for SMA patients. Markers assessed in the current study corroborate the premise of systemic disease manifestation and highlight the importance of close monitoring for emergence of disease phenotypes beyond muscle denervation.

In summary, this case series analysis study highlights the importance of early intervention with gene therapy and other molecular treatments for SMA and the need for close monitoring of treatment responses beyond muscle denervation in the real world clinical setting.

## Data Availability Statement

All data relevant to this study are contained within the manuscript.

## Acknowledgements

We are grateful to all the patients and families who participated in this study.

## Author Contributions

K.J.S. and C.R.R.A. directed the research project. R.G.S., S.D.L., E.A.A., E.R., and K.J.S. collected clinical data. R.G.S., L.L.H., S.D.L., E.A.A., E.R., K.J.S., and C.R.R.A. analyzed the data and participated in the data interpretation. L.L.H., K.J.S. and C.R.R.A. drafted the manuscript. All authors reviewed and approved the final manuscript.

## Funding

C.R.R.A. received a fellowship from the MGH ECOR. K.J.S. was funded by NIH NICHD R01HD054599, NIH NINDS R21NS108015, Biogen, and Cure SMA.

### Ethical Approval

Ethical approval and written informed parental consent were obtained from all participants under Institutional Ethics Review Boards at the Massachusetts General Hospital (protocol 2016P000469).

## Figure Legends

**Supplementary Information**. The Excel table consists of a comprehensive set of safety lab data for each patient. Reference values for each lab measurement were annotated from Mayo Clinic and UCSF Benioff Children’s Hospital.

