## Supplemental Table 1 for "Early Life Safety Profiling of Gene Therapy for Spinal Muscular Atrophy: A Case Series Analysis"

| ID | LAB TYPE | DAYS POST DOSE | MEASUREMENT |
|---|---|---|---|
| | Albumin (g/dL) | -855 | 3.4 |
| | | -696 | 4.1 |
| | | -582 | 4.2 |
| | | -100 | 4.3 |
| | | -11 | 4 |
| | | 27 | 3.5 |
| | | 54 | 4.7 |
| | | 108 | 4.4 |
| | | 139 | 4.7 |
| | | 216 | 4.2 |
| | | 424 | 4.7 |
| | Gamma Glutamyl Transferase (GGT) (U/L) | -100 | 9 |
| | | -11 | 10 |
| | | 27 | 9 |
| | | 54 | 72 |
| | | 108 | 15 |
| | | 139 | 7 |
| | | 424 | 10 |
| | Troponin T-hs Gen5 (ng/L) | -100 | 69 |
| | | -11 | 26 |
| | | 27 | 19 |
| | | 54 | 22 |
| | | 108 | 61 |
| | | 139 | 52 |
| | HGB | -861 | 9.7 |
| | | -855 | 8.2 |
| | | -850 | 10 |
| | | -841 | 10.6 |
| | | -793 | 11.5 |
| | | -696 | 11.4 |
| | | -582 | 12.1 |
| | | -452 | 11.8 |
| | | -100 | 12.2 |
| | | -11 | 12.1 |
| | | 27 | 12.2 |
| | | 54 | 14.8 |
| | | 108 | 13.5 |
| | | 139 | 12.7 |
| | | 216 | 9.5 |
| | | 381 | 12.6 |

| | | | |
|---|---|---|---|
| 9 | | 424 | 12.7 |
| | Nucleated RBCs. Automated (# per 100 WBCs) | -855 | 0 |
| | | -841 | 0 |
| | | -793 | 0 |
| | | -696 | 0 |
| | | -452 | 0 |
| | | -100 | 0 |
| | | -11 | 0.2 |
| | | 381 | 0.02 |
| | Platelet Count (1000 / uL) | -861 | 791 |
| | | -855 | 499 |
| | | -850 | 422 |
| | | -841 | 515 |
| | | -793 | 338 |
| | | -696 | 373 |
| | | -582 | 374 |
| | | -452 | 533 |
| | | -100 | 453 |
| | | -11 | 358 |
| | | 27 | 355 |
| | | 54 | 156 |
| | | 108 | 409 |
| | | 139 | 430 |
| | | 216 | 541 |
| | | 381 | 243 |
| | | 424 | 415 |
| | WBC 10^3/uL | -861 | 19 |
| | | -855 | 15.7 |
| | | -850 | 7.3 |
| | | -841 | 4.96 |
| | | -793 | 6.6 |
| | | -696 | 9.38 |
| | | -582 | 11.31 |
| | | -452 | 14.83 |
| | | -100 | 10.29 |
| | | -11 | 10.63 |
| | | 27 | 12.04 |
| | | 54 | 11.51 |
| | | 108 | 7.92 |
| | | 139 | 7.23 |
| | | 216 | 13.11 |

| | | | |
|---|---|---|---|
| | | 381 | 7.87 |
| | | 424 | 7.8 |
| 1 | Albumin (g/dL) | -9 | 3.7 |
| | | 6 | 3.7 |
| | | 12 | 3.9 |
| | | 29 | 4.5 |
| | | 63 | 4.1 |
| | | 75 | 4.2 |
| | | 149 | 4.4 |
| | | 230 | 4.8 |
| | | 336 | 4.7 |
| | | 426 | 4.8 |
| | | 523 | 4.8 |
| | Gamma Glutamyl Transferase (GGT) (U/L) | -9 | |
| | | 6 | 66 |
| | | 12 | 52 |
| | | 29 | 38 |
| | | 29 | 38 |
| | | 63 | 38 |
| | | 75 | 28 |
| | | 230 | 18 |
| | | 336 | 18 |
| | | 426 | 17 |
| | | 523 | 14 |
| | HGB | -9 | 11.6 |
| | | 6 | 13.2 |
| | | 29 | 11.1 |
| | | 63 | 11.8 |
| | | 75 | 11.6 |
| | | 230 | 11.4 |
| | | 336 | 10.5 |
| | | 426 | 10.8 |
| | | 523 | 12 |
| | Nucleated RBCs. Automated (# per 100 WBCs) | 6 | 0.3 |
| | | 63 | 0 |
| | | 75 | 0.2 |
| | | 230 | 0 |
| | | 426 | 0 |
| | | 523 | 0 |
| | | -9 | 284 |
| | | 6 | 385 |

| | | | |
|---|---|---|---|
| | Platelet Count (1000 / uL) | 29 | 350 |
| | | 63 | 400 |
| | | 75 | 404 |
| | | 230 | 497 |
| | | 336 | 434 |
| | | 426 | 449 |
| | | 523 | 346 |
| | WBC 10^3/uL | -9 | 6.88 |
| | | 6 | 7.55 |
| | | 29 | 6.58 |
| | | 63 | 7.44 |
| | | 75 | 8.09 |
| | | 230 | 13.16 |
| | | 336 | 10.03 |
| | | 426 | 10.43 |
| | | 523 | 10.47 |
| 12 | Albumin (g/dL) | 0 | 4 |
| | | 62 | 4.1 |
| | | 83 | 4.3 |
| | | 167 | 4.2 |
| | | 280 | 4.6 |
| | | 392 | 4.7 |
| | | 504 | 4.8 |
| | | 708 | 5.1 |
| | | 886 | 4.9 |
| | Gamma Glutamyl Transferase (GGT) (U/L) | 62 | 46 |
| | | 83 | 33 |
| | | 167 | 11 |
| | | 280 | 8 |
| | | 392 | 13 |
| | | 504 | 11 |
| | | 708 | 8 |
| | Troponin T-hs Gen5 (ng/L) | 0 | 82 |
| | | 62 | 42 |
| | | 83 | 45 |
| | | 167 | 90 |
| | | 280 | 73 |
| | | 392 | 66 |
| | | 504 | 59 |

| | | |
|---|---|---|
| | 886 | 41 |
| HGB | 62 | 12.3 |
| | 167 | 11 |
| | 280 | 11.7 |
| | 392 | 12.3 |
| | 504 | 13 |
| | 708 | 13.3 |
| | 886 | 14 |
| Nucleated RBCs. Automated (# per 100 WBCs) | 62 | 0 |
| | 392 | 0 |
| Platelet Count (1000 / uL) | 62 | 354 |
| | 167 | 289 |
| | 280 | 423 |
| | 392 | 308 |
| | 504 | 378 |
| | 708 | 389 |
| | 886 | 329 |
| WBC 10^3/uL | 62 | 6.15 |
| | 167 | 5.71 |
| | 280 | 10.23 |
| | 392 | 8.54 |
| | 504 | 10.85 |
| | 708 | 9 |
| | 886 | 7.02 |
| Albumin (g/dL) | -3 | 4.3 |
| | 20 | 3.8 |
| | 41 | 4.1 |
| | 55 | 4.2 |
| | 68 | 4.7 |
| | 88 | 4.5 |
| | 171 | 4.6 |
| | 262 | 4.4 |
| | 353 | 4.4 |
| Gamma Glutamyl Transferase (GGT) (U/L) | -3 | 189 |
| | -3 | 189 |
| | 20 | 55 |
| | 41 | 49 |
| | 55 | 76 |

| | | | |
|---|---|---|---|
| | Gamma Glutamyl Transferase (GGT) (U/L) | 68 | 62 |
| | | 88 | 38 |
| | | 171 | 18 |
| | | 262 | 12 |
| | | 353 | 12 |
| | Troponin T-hs Gen5 (ng/L) | 595 | 5 |
| 3 | HGB | -3 | 15 |
| | | 20 | 12.4 |
| | | 55 | 13.3 |
| | | 88 | 12.8 |
| | | 171 | 9.7 |
| | | 262 | 13.7 |
| | | 353 | 11.8 |
| | Nucleated RBCs. Automated (# per 100 WBCs) | -3 | 0 |
| | | 171 | 114 |
| | | 262 | 100 |
| | | 353 | 128 |
| | Platelet Count (1000 / uL) | -3 | 491 |
| | | 20 | 689 |
| | | 55 | 628 |
| | | 88 | 693 |
| | | 171 | 730 |
| | | 262 | 422 |
| | | 353 | 365 |
| | WBC 10^3/uL | -3 | 15.71 |
| | | 20 | 12.73 |
| | | 55 | 10.41 |
| | | 88 | 12.71 |
| | | 171 | 10.4 |
| | | 262 | 12.97 |
| | | 353 | 13.5 |
| | Albumin (g/dL) | -49 | 4.3 |
| | | 0 | 4.6 |
| | | 12 | 4 |
| | | 89 | 4.7 |
| | | 152 | 4.7 |
| | | 236 | 4.8 |
| | Gamma Glutamyl Transferase (GGT) (U/L) | -49 | 26 |
| | | 0 | 23 |
| | | 12 | 15 |
| | | 152 | 12 |

| | | | |
|---|---|---|---|
| 6 | | 236 | 11 |
| | Troponin T-hs Gen5 (ng/L) | -49 | 28 |
| | | 0 | 20 |
| | | 236 | 7 |
| | HGB | -49 | 9 |
| | | 0 | 8.2 |
| | | 1 | 8.3 |
| | | 6 | 8.9 |
| | | 89 | 12.7 |
| | | 236 | 14.3 |
| | Platelet Count (1000 / uL) | -49 | 914 |
| | | 0 | 608 |
| | | 1 | 653 |
| | | 6 | 583 |
| | | 89 | 446 |
| | | 236 | 422 |
| | WBC 10^3/uL | -49 | 10.82 |
| | | 0 | 9.95 |
| | | 1 | 10.3 |
| | | 6 | 6.7 |
| | | 89 | 5.61 |
| | | 236 | 10.26 |
| 4 | Albumin (g/dL) | 0 | 4 |
| | | 39 | 4.4 |
| | | 60 | 4.2 |
| | | 80 | 4 |
| | | 283 | 3.7 |
| | | 453 | 4.3 |
| | Gamma Glutamyl Transferase (GGT) (U/L) | 0 | 29 |
| | | 39 | 18 |
| | | 60 | 16 |
| | | 80 | 14 |
| | | 453 | 11 |
| | Troponin T-hs Gen5 (ng/L) | 0 | 29 |
| | | 39 | 11 |
| | | 80 | 23 |
| | | 283 | 7 |
| | | 453 | **???** |
| | | 0 | 9.8 |

| | | |
|---|---|---|
| HGB | 60 | 12.1 |
| | 80 | 12.1 |
| | 283 | 11 |
| | 453 | 11.8 |
| Nucleated RBCs. Automated (# per 100 WBCs) | 0 | 0 |
| | 80 | 109 |
| Platelet Count (1000 / uL) | 0 | 446 |
| | 60 | 346 |
| | 80 | 395 |
| | 283 | 362 |
| | 453 | 319 |
| WBC 10^3/uL | 0 | 7.06 |
| | 60 | 7.1 |
| | 80 | 11.42 |
| | 283 | 11.6 |
| | 453 | 8.48 |
| Albumin (g/dL) | -5 | 4.5 |
| | 14 | 4.1 |
| | 21 | 4.4 |
| | 33 | 4.1 |
| | 41 | 4.4 |
| | 155 | 4.4 |
| | 288 | 4.9 |
| | 517 | 4.9 |
| Gamma Glutamyl Transferase (GGT) (U/L) | -5 | 30 |
| | 14 | 50 |
| | 21 | 30 |
| | 33 | 5 |
| | 41 | 31 |
| | 155 | 12 |
| | 288 | 13 |
| | 517 | 10 |
| Troponin T-hs Gen5 (ng/L) | -5 | 39 |
| | 14 | 39 |
| | 21 | 40 |
| | 33 | 82 |
| | 41 | 33 |
| | 155 | 51 |
| | 288 | 64 |

10

| | | |
|---|---|---|
| HGB | -13 | 12.3 |
| | -5 | 11.7 |
| | 14 | 11.2 |
| | 21 | 11.7 |
| | 33 | 10.8 |
| | 41 | 12.1 |
| | 155 | 10.6 |
| | 288 | 11.1 |
| | 517 | 12.9 |
| Nucleated RBCs. Automated (# per 100 WBCs) | -5 | 0 |
| | 14 | 100 |
| | 33 | 0 |
| | 41 | 0 |
| Platelet Count (1000 / uL) | -13 | 478 |
| | -5 | 655 |
| | 14 | 871 |
| | 21 | 281 |
| | 33 | 326 |
| | 41 | 541 |
| | 155 | 489 |
| | 288 | 318 |
| WBC 10^3/uL | -13 | 11.3 |
| | -5 | 15.43 |
| | 14 | 7.58 |
| | 21 | 6.04 |
| | 33 | 16.51 |
| | 41 | 10.32 |
| | 155 | 9.74 |
| | 288 | 14.02 |
| | 517 | 14.72 |
| Albumin (g/dL) | -63 | 3.9 |
| | -8 | 4.3 |
| | -1 | 4.1 |
| | 84 | 4.3 |
| | 195 | 4.2 |
| | 321 | 4.6 |
| | 446 | 4.7 |
| | -8 | 50 |
| | -1 | 40 |

| | | | |
|---|---|---|---|
| 11 | Gamma Glutamyl Transferase (GGT) (U/L) | 84 | 18 |
| | | 321 | 6 |
| | | 446 | 13 |
| | Troponin T-hs Gen5 (ng/L) | -8 | 42 |
| | | -1 | 39 |
| | | 195 | 60 |
| | HGB | -93 | 13 |
| | | -63 | 11.7 |
| | | -1 | 13.4 |
| | | 84 | 13.2 |
| | | 195 | 10.9 |
| | | 321 | 12.5 |
| | | 446 | 12.4 |
| | Nucleated RBCs. Automated (# per 100 WBCs) | -93 | 0 |
| | | -63 | 113 |
| | | -1 | 0 |
| | | 84 | 0 |
| | | 195 | 0 |
| | Platelet Count (1000 / uL) | -93 | 677 |
| | | -63 | 562 |
| | | -1 | 160 |
| | | 84 | 316 |
| | | 195 | 353 |
| | | 321 | 357 |
| | | 446 | 476 |
| | WBC 10^3/uL | -93 | 8.04 |
| | | -63 | 9.87 |
| | | -1 | 8.8 |
| | | 84 | 6.75 |
| | | 195 | 13.5 |
| | | 321 | 10.25 |
| | | 446 | 8.5 |
| | Albumin (g/dL) | | |
| | | -1 | 4.3 |
| | | 82 | 4.4 |
| | | 187 | 4.2 |
| | | 397 | 4.2 |
| | Gamma Glutamyl Transferase (GGT) (U/L) | -1 | 13 |
| | | 82 | 15 |
| | | 187 | 16 |

| | | | |
|---|---|---|---|
| 7 | | 397 | 7 |
| | Troponin T-hs Gen5 (ng/L) | -1 | 13 |
| | | 82 | 7 |
| | | 187 | 6 |
| | HGB | -1 | 9.5 |
| | | 82 | 12 |
| | | 187 | 10.4 |
| | | 397 | 10.7 |
| | Nucleated RBCs. Automated (# per 100 WBCs) | -1 | 0 |
| | | 82 | 0 |
| | | 187 | 0 |
| | | 397 | 0 |
| | Platelet Count (1000 / uL) | -1 | 547 |
| | | 82 | 407 |
| | | 187 | 363 |
| | WBC 10^3/uL | -1 | 12.68 |
| | | 82 | 9.56 |
| | | 187 | 9.34 |
| | | 397 | 6.02 |
| 8 | Albumin (g/dL) | 8 | 4.6 |
| | | 36 | 4.6 |
| | | 57 | 4.3 |
| | | 85 | 4.4 |
| | | 311 | 4.5 |
| | | 436 | 4.5 |
| | Gamma Glutamyl Transferase (GGT) (U/L) | 8 | 18 |
| | | 36 | 34 |
| | | 57 | 32 |
| | | 85 | 20 |
| | Troponin T-hs Gen5 (ng/L) | 8 | 21 |
| | | 36 | 8 |
| | | 57 | 11 |
| | | 85 | 13 |
| | HGB | 8 | 11.7 |
| | | 36 | 12.5 |
| | | 57 | 12.4 |
| | | 85 | 12.7 |
| | | 311 | 11.5 |
| | | 436 | 11.9 |

| | | | |
|---|---|---|---|
| | Nucleated RBCs. Automated (# per 100 WBCs) | 8 | 0 |
| | | 85 | 0 |
| | Platelet Count (1000 / uL) | 8 | 181 |
| | | 36 | 333 |
| | | 57 | 333 |
| | | 85 | 246 |
| | | 311 | 266 |
| | | 436 | 282 |
| | WBC 10^3/uL | 8 | 3.96 |
| | | 36 | 7.48 |
| | | 57 | 7.16 |
| | | 85 | 6.43 |
| | | 311 | 5.84 |
| | | 436 | 6.6 |
| 13 | Albumin (g/dL) | -11 | 4 |
| | | 17 | 4.1 |
| | | 39 | 3.3 |
| | | 62 | 4.5 |
| | | 97 | 4.5 |
| | | 143 | 4.1 |
| | | 360 | 4.7 |
| | Creatinine Kinase (CK): (U/L) | 360 | 141 |
| | Gamma Glutamyl Transferase (GGT) (U/L) | -11 | 22 |
| | | 17 | 19 |
| | | 97 | 8 |
| | | 360 | 4 |
| | HGB | -23 | 12.5 |
| | | -11 | 10.3 |
| | | 17 | 11 |
| | | 39 | 9.4 |
| | | 62 | 13.8 |
| | | 97 | 12.1 |
| | | 143 | 10.2 |
| | | 360 | 10.8 |
| | Nucleated RBCs. Automated (# per 100 WBCs) | -23 | 0 |
| | | 17 | 0 |
| | | 62 | 0 |
| | | 143 | 0 |
| | | -23 | 629 |

| | | | |
|---|---|---|---|
| | Platelet Count (1000 / uL) | -11 | 417 |
| | | 17 | 662 |
| | | 39 | 799 |
| | | 62 | 516 |
| | | 97 | 407 |
| | | 143 | 267 |
| | | 360 | 431 |
| | WBC 10^3/uL | -23 | 11.21 |
| | | -11 | 7.07 |
| | | 17 | 7.46 |
| | | 39 | 11.48 |
| | | 62 | 11.9 |
| | | 97 | 11.15 |
| | | 143 | 8.23 |
| | | 360 | 10.86 |
| 5 | Albumin (g/dL) | | 3.7 |
| | | 80 | |
| | | 153 | 4.2 |
| | | 230 | 4.5 |
| | | 426 | 4.3 |
| | | 519 | 4.2 |
| | Gamma Glutamyl Transferase (GGT) (U/L) | 80 | 12 |
| | | 153 | 13 |
| | | 230 | 11 |
| | | 426 | 7 |
| | | 519 | 7 |
| | Troponin T-hs Gen5 (ng/L) | 80 | 25 |
| | | 153 | 11 |
| | | 230 | 8 |
| | | 426 | 7 |
| | | 519 | **???** |
| | HGB | 80 | 11.4 |
| | | 153 | 11.5 |
| | | 230 | 12 |
| | | 426 | 11.2 |
| | | 519 | 11.2 |
| | Platelet Count (1000 / uL) | 80 | 609 |
| | | 153 | 605 |
| | | 230 | 347 |
| | | 519 | 406 |

| | | | |
|---|---|---|---|
| | WBC 10^3/uL | 80 | 15.58 |
| | | 153 | 19.85 |
| | | 230 | 17.72 |
| | | 426 | 13.66 |
| | | 519 | 13.44 |
| 2 | Albumin (g/dL) | 81 | 4.2 |
| | | 116 | 3.9 |
| | | 257 | 4.4 |
| | Gamma Glutamyl Transferase (GGT) (U/L) | 257 | 10 |
| | HGB | 81 | 14.1 |
| | | 257 | 14.5 |
| | Platelet Count (1000 / uL) | 81 | 371 |
| | | 257 | 378 |
| | WBC 10^3/uL | 81 | 10.8 |
| | | 257 | 11.2 |
| | Albumin (g/dL) | -60 | 4 |
| | | -5 | 4.4 |
| | | 8 | 4.2 |
| | | 25 | 4 |
| | | 43 | 4.4 |
| | | 57 | 4.3 |
| | | 71 | 4.5 |
| | | 93 | 4.5 |
| | | 218 | 4.8 |
| | Gamma Glutamyl Transferase (GGT) (U/L) | -5 | 24 |
| | | 8 | 23 |
| | | 25 | 14 |
| | | 43 | 11 |
| | | 57 | 17 |
| | | 71 | 16 |
| | | 93 | 19 |
| | Troponin T-hs Gen5 (ng/L) | -60 | 38 |
| | | -5 | 36 |
| | | 25 | 68 |
| | | 43 | 46 |
| | | 57 | 25 |
| | | 93 | 25 |
| | | -32 | 9.1 |
| | | -5 | 10 |

| | | | |
|---|---|---|---|
| 14 | HGB | 8 | 10.9 |
| | | 25 | 11.1 |
| | | 43 | 11.1 |
| | | 57 | 11 |
| | | 93 | 12 |
| | | 218 | 12 |
| | | 272 | 11.2 |
| | Nucleated RBCs. Automated (# per 100 WBCs) | 8 | 0 |
| | | 25 | 0 |
| | | 43 | 0 |
| | | 218 | 0 |
| | Platelet Count (1000 / uL) | -32 | 613 |
| | | -5 | 467 |
| | | 8 | 199 |
| | | 25 | 415 |
| | | 43 | 377 |
| | | 57 | 389 |
| | | 93 | 365 |
| | | 218 | 356 |
| | | 272 | 399 |
| | WBC 10^3/uL | -32 | 6.43 |
| | | -5 | 5.06 |
| | | 8 | 7.91 |
| | | 25 | 6.75 |
| | | 43 | 7.04 |
| | | 57 | 5.87 |
| | | 93 | 5.91 |
| | | 218 | 8.18 |
| | | 272 | 7.8 |

| POST-DOSE RANGE | NORMAL RANGE |
|---|---|
| 3.5 - 4.7 | 3.3 - 5.0 |
| 7 - 72 | <21 (12m-6y) |
| 19 - 61 | < or =10 (F) |
| 9.5 - 14.8 | 10.2-12.7 (6-35m &F)<br>11.4-14.3 (3-5y & F) |

Measurements ou

| | |
|---|---|
| 0.02 - 0.02 | 0 |
| 156 - 541 | 189-394 (24-35m & F)<br>187-445 (3-5y) |
| 7.23 - 13.11 | 4.9-13.2 (24-35m & F)<br>4.4-12.9 (3-5y) |

| | |
|---|---|
| 3.7 - 4.8 | 3.3 - 5.0 |
| 14 - 66 | <178 (0-11m)<br><21 (12m-6y) |
| 10.5 - 13.2 | 9.2-11.4 (5-7w & F)<br>9.9-12.4 (8w-5m & F)<br>10.2-12.7 (6-35m & F) |
| 0 - 0.3 | 0 |
| | 331-597 (5-7w & F) |

| | |
|---|---|
| 346 - 497 | 247-580 (8w-5m & F)<br>214-459 (6-23m & F) |
| 6.58 - 13.16 | 7.1-14.7 (5-7w & F)<br>6.0-13.3 (8w-5m & F)<br>6.5-13.0 (6-23m & F) |
| 4.1 - 5.1 | 3.3 - 5.0 |
| 8 - 46 | <178 (0-11m)<br><21 (12m-6y) |
| 41 - 90 | < or =15 (M) |

| | |
|---|---|
| 11 - 14 | 10.1-12.5 (6-23m & M)<br>10.2-12.7 (24-35m & M) |
| 0 | 0 |
| 289 - 423 | 206-445 (6-23m & M)<br>202-403 (24-35m & M) |
| 5.71 - 10.85 | 6.0-13.5 (6-23m & M)<br>5.1-13.4 (24-35m & M) |
| 3.8 - 4.7 | 3.3 - 5.0 |
| 12 - 76 | <178 (0-11m) |

| | |
|---|---|
| 12 - 7[illegible] | <21 (12m-6y) |
| 5 | <21 (12m-6y) |
| 9.7 - 13.7 | 9.2-11.4 (5-7w & F)<br>9.9-12.4 (8w-5m & F)<br>10.2-12.7 (6-35m & F) |
| 100 - 128 | 0 |
| 365 - 730 | 331-597 (5-7w & F)<br>247-580 (8w-5m & F)<br>214-459 (6-23m & F) |
| 10.4 - 13.5 | 7.1-14.7 (5-7w & F)<br>6.0-13.3 (8w-5m & F)<br>6.5-13.0 (6-23m & F) |
| 4 - 4.8 | 3.3 - 5.0 |
| 11 - 15 | <178 (0-11m)<br><21 (12m-6y) |

| | |
|---|---|
| 7 | < or =15 (M) |
| 8.3 - 14.3 | 10.1-12.5 (6m-23m & M) |
| 422 - 653 | 206-445 (6-23m & M) |
| 5.61 - 10.3 | 6.0-13.5 (6-23m & M) |
| 3.7 - 4.4 | 3.3 - 5.0 |
| 11 - 18 | <178 (0-11m)<br><21 (12m-6y) |
| 7 - 23 | < or =10 (F) |

| | |
|---|---|
| 11 - 12.1 | 9.9-12.4 (8w-5m & F)<br>10.2-12.7 (6-35m & F) |
| 109 | 0 |
| 319 - 395 | 247-580 (8w-5m & F)<br>214-459 (6-23m & F) |
| 7.1 - 11.6 | 6.0-13.3 (8w-5m & F)<br>6.5-13.0 (6-23m & F) |
| 4.1 - 4.9 | 3.3 - 5.0 |
| 5 - 50 | <178 (0-11m)<br><21 (12m-6y) |
| 33 - 82 | < or =10 (F) |

| | |
|---|---|
| 10.6 - 12.9 | 9.9-12.4 (8w-5m & F)<br>10.2-12.7 (6-35m & F) |
| 0-100 | 0 |
| 281 - 871 | 247-580 (8w-5m & F)<br>214-459 (6-23m & F) |
| 6.04 - 16.51 | 6.0-13.3 (8w-5m & F)<br>6.5-13.0 (6-23m & F) |
| 4.2 - 4.7 | 3.3 - 5.0 |
| | <178 (0-11m) |

| | |
|---|---|
| 6 - 18 | <21 (12m-6y) |
| 60 | < or =10 (F) |
| 10.9 - 13.2 | 10.2-12.7 (6-35m & F) |
| 0 | 0 |
| 316 - 476 | 214-459 (6-23m & F) |
| 6.75 - 13.5 | 6.5-13.0 (6-23m & F) |
| 4.2 - 4.4 | 3.3 - 5.0 |
| 7 - 16 | <178 (0-11m)<br><21 (12m-6y) |

| | |
|---|---|
| | <21 (12m-6y) |
| 6 - 7 | < or =10 (F) |
| 10.4 - 12 | 10.2-12.7 (6-35m & F) |
| 0 | 0 |
| 363 - 407 | 214-459 (6-23m & F) |
| 6.02 - 9.56 | 6.5-13.0 (6-23m & F) |
| 4.3 - 4.6 | 3.3 - 5.0 |
| 18 - 34 | <178 (0-11m) |
| 8 - 21 | < or =15 (M) |
| 11.5 - 12.7 | 10.1-12.5 (6m-23m & M) |

| | |
|---|---|
| 0 | 0 |
| 181 - 333 | 206-445 (6-23m & M) |
| 3.96 - 7.48 | 6.0-13.5 (6-23m & M) |
| 3.3 - 4.7 | 3.3 - 5.0 |
| 141 | 39-308 (M) |
| 4 - 19 | <178 (0-11m)<br><21 (12m-6y) |
| 9.4 - 13.8 | 9.6-12.4 (8w-5m & M)<br>10.1-12.5 (6m-23m & M) |
| 0 | 0 |

| | |
|---|---|
| 267 - 799 | 244-529 (8w-5m & M)<br>206-445 (6-23m & M) |
| 7.46 - 11.9 | 6.5-13.3 (8w-5m & M)<br>6.0-13.5 (6-23m & M) |
| 3.7 - 4.5 | 3.3 - 5.0 |
| 7 - 13 | <178 (0-11m)<br><21 (12m-6y) |
| 7 - 25 | < or =10 (F) |
| 11.2 - 12 | 9.9-12.4 (8w-5m & F)<br>10.2-12.7 (6-35m & F) |
| 347 - 609 | 247-580 (8w-5m & F)<br>214-459 (6-23m & F) |

| | |
|---|---|
| 13.44 - 19.85 | 6.0-13.3 (8w-5m & F)<br>6.5-13.0 (6-23m & F) |
| 3.9 - 4.4 | 3.3 - 5.0 |
| 10 | <178 (0-11m) |
| 14.1 - 14.5 | 9.6-12.4 (8w-5m & M)<br>10.1-12.5 (6m-23m & M) |
| 371 - 378 | 244-529 (8w-5m & M)<br>206-445 (6-23m & M) |
| 10.8 - 11.2 | 6.5-13.3 (8w-5m & M)<br>6.0-13.5 (6-23m & M) |
| 4 - 4.8 | 3.3 - 5.0 |
| 11 - 23 | <178 (0-11m) |
| 25 - 68 | < or =15 (M) |

| | |
|---|---|
| 10.9 - 12 | 9.6-12.4 (8w-5m & M)<br>10.1-12.5 (6m-23m & M) |
| 0 | 0 |
| 199 - 415 | 244-529 (8w-5m & M)<br>206-445 (6-23m & M) |
| 5.87 - 8.18 | 6.5-13.3 (8w-5m & M)<br>6.0-13.5 (6-23m & M) |

DA

ıt of normal range post injection are highlighted in orange.

AYS POST DOSE

| Source | Mayo Clinic Laboratories<br>(Date accessed: 8/19/2024) | UCSF Benioff Children's Hospital<br>(Date accessed: 8/19/2024) |
|---|---|---|
| **Albumin** | 3.5-5.0 g/dL (> or =12 mo) | 3.4-5.4 g/dL |
| **GGT** | <178 U/L (0-11 mo)<br><21 U/L (12mo-6y) | 5-40 U/L |
| **Troponin** | < or =15 ng/L (M)<br>< or =10 ng/L (F) | “Normally so low they cannot be detected” |
| **HGB** | MALES: g/dL<br>13.9-19.1 (0-14d)<br>10.0-15.3 (15d-4w)<br>8.9-12.7 (5w-7w)<br>9.6-12.4 (8w-5m)<br>10.1-12.5 (6m-23m)<br>10.2-12.7 (24m-35m)<br>11.4-14.3 (3-5y)<br>FEMALES:<br>13.4-20.0 (0-14d)<br>10.8-14.6 (15d-4w)<br>9.2-11.4 (5-7w)<br>9.9-12.4 (8w-5m)<br>10.2-12.7 (6-35m)<br>11.4-14.3 (3-5y) | MALE: 13.8-17.2 gm/dL<br>FEMALE: 12.1-15.1 gm/dL |
| **Nucleated RBCs** | 0 NRBCs/100 WBC | N/A |
| **Platelet count** | MALES:<br>218-419 (0-14d)<br>248-586 (15d-4w)<br>229-562 (5-7w)<br>244-529 (8w-5m)<br>206-445 (6-23m)<br>202-403 (24-35m)<br>187-445 (3-5y)<br>FEMALES:<br>144-449 (0-14d)<br>279-571 (15d-4w)<br>331-597 (5-7w)<br>247-580 (8w-5m)<br>214-459 (6-23m)<br>189-394 (24-35m)<br>187-445 (3-5y) | 150,000-450,000/dL |
| | MALES: 10(9)/L<br>8.0-15.4 (0-14d)<br>7.8-15.9 (15d-4w)<br>8.1-15.0 (5-7w) | |

| | | |
|---|---|---|
| **WBC** | 6.5-13.3 (8w-5m)<br>6.0-13.5 (6-23m)<br>5.1-13.4 (24-35m)<br>4.4-12.9 (3-5y)<br>FEMALES:<br>8.2-14.6 (0-14d)<br>15 days-4 weeks: 8.4-14.4 (15d-4w)<br>7.1-14.7 (5-7w)<br>6.0-13.3 (8w-5m)<br>6.5-13.0 (6-23m)<br>4.9-13.2 (24-35m)<br>4.4-12.9 (3-5y) | 4,500-10,000 cells/mcL |
| **Creatine Kinase** | MALES: 39-308 U/L (> 3m)<br>FEMALES: 26-192 U/L (> 3m) | 10-120 mcg/L |
